# Psychometric Properties of a New Stress-Related Personality Scale: The Virtual Inventory of Behavior and Emotions (VIBE)

**DOI:** 10.1101/2021.12.08.21267487

**Authors:** David C. Rettew, Dustin Freckleton, Nicholas Schork, Arnulf Graf, Zoe Morrell, Lucy Lincoln, Susan Smalley, James J. Hudziak

**Author notes:** Corresponding Author: David Rettew MD, Lane County Behavioral Health, 2411 Martin Luther King Jr Blvd, Eugene, OR 97401.

## Abstract

**Background:** Personality traits are important factors with regard to the tendency to experience and response to stress. This study introduces and tests a new stress-related personality scale called the Virtual Inventory of Behavior and Emotions (VIBE).

**Methods:** Two samples totaling 5512 individuals (with 66% between the ages of 18 and 34) completed the VIBE along with other measures of personality, stress, mood, and well-being. Results: Exploratory factor analyses revealed a four-factor structure for the instrument with dimensions labelled: 1) stressed; 2) energetic; 3) social; and 4) disciplined. Confirmatory factor analytic procedures on the final 23-item version showed good psychometric properties and data fit while machine learning analyses demonstrated the VIBE’s ability to distinguish between groups with similar patterns of response. Strong convergent validity was suggested through robust correlations between the dimensions of the VIBE and other established rating scales.

**Conclusion:** Overall, the data suggest that the VIBE is a promising tool to help advance understanding of the relations between stress, personality, and related constructs.

## INTRODUCTION

Research on personality traits has increasingly demonstrated an important role for personality traits in both physical and emotional-behavioral health. Wide ranging studies have specifically shown that particular personality dimensions are significantly related to overall well-being, the probability of developing a psychiatric disorder, how long one can expect to live, and financial success, among other outcomes.^1-5^ The effect sizes of many of these associations have been found to be comparable to other important domains such as intelligence and socioeconomic status.^6^ Indeed, various personality traits related to the ability to self-regulate and pursue goals have been judged as “the most important psychological asset a social actor can have.”^4^

Studies have also revealed that personality traits can have profound impacts on a person’s tendency to experience more frequent and intense levels of stress and how that stress is expressed through a number of physiological parameters.^7^ For example, a study of students in Switzerland exposed to a campus shooting alarm that triggered a massive police response found that the trait of neuroticism was associated with heightened panic and trauma-related stress, while agreeableness and conscientiousness were found to be related to the stress-related coping strategy of seeking out social support.^8^ Recent studies of college students assessed during the COVID pandemic revealed strong associations not only between several personality dimensions (higher levels of neuroticism and lower levels of extraversion) and perceived stress, but also with the employment of emotion versus problem-based coping strategies.^9,10^ These associations may be mediated through more specific processes such as perceiving higher levels of threat and having less confidence in one’s ability to effectively reduce that threat.^11^

Over the past thirty years, a number of studies have reported substantial genetic influences (0.42-0.52%) on the development of personality. However, these same percentages also indicate that environmental factors contribute as much or more to the variance within specific traits^12^ with interactions between genetic and environmental factors leading to both stability and change in an individual’s personality over time.^13^ Among specific environmental factors, stress has been shown to have direct impacts on personality development.^14^ For these reasons, we were interested in including stress-related variables in our study of personality to help examine these bi-directional influences.

There are a number of instruments and rating scales designed to assess personality and related constructs such as temperament and character. Despite a variety of methods and a diversity of theoretical foundations that underlie them, many of the most commonly-used scales have settled on a relatively small number of core traits related to predispositions toward negative and positive emotions, sociability, activity level, sensation seeking, regulatory abilities, and goal-directed behavior.^15-18^ While these tools have provided a wealth of important data on personality traits and their relevance to a number of different areas, many personality assessment instruments were written decades ago and may no longer be as applicable to the younger Millennial and Gen Z generations. The Big Five Inventory, for example, includes an item about feeling “blue” while the Temperament and Character Inventory uses terms such as “eager beaver.”^16,19^ Such terms are no longer commonly used in everyday language among younger individuals and could lead to confusion and potentially even reduced engagement in the testing process. Many of these traditional instruments also can be long and time consuming in an era where people increasingly struggle to read fully through online content.^20^ Finally, most personality instruments were not designed to look specifically at the relations between various personality traits and stress.

For these reasons, we decided to undertake the development of a new stress-related personality scale that could be used in conjunction with other instruments to assess how stress is related to physiological parameters and predict who may be more vulnerable to experiencing stress. Our ultimate goal was to design an instrument that could be easily given online or completed on a smartphone in a short amount of time. We also wanted to use language for the individual items that reflects current speech and to build items that incorporate modern life, such as the ubiquitous use of social media and related technologies. At the same time, it was also important to have an instrument that was psychometrically valid and that accurately reflected the dimensions it was designed to capture.

The present study describes our initial investigations into this new stress-related personality instrument, called the Virtual Inventory of Behavior and Emotions (VIBE). The hypothesis underlying our work was that we would be able to develop an instrument that not only would be concise but show strong psychometric properties and exhibit significant associations with both established personality scales and with validated measures of stress, mood, and well-being.

## METHODS

### Initial VIBE Construction

The original items of the VIBE were created by several of the coauthors (DR, JH, DF). These items were designed to be loosely based on the “Big Five” structure of personality.^21^ Emphasis was placed on creating statements (i.e., items) that would be particularly relevant to stress proneness and resilience that used language related to behaviors and feelings relevant for younger adults. Out of an initial 67 statements to which a respondent would rate their level of agreement, a battery of 42 questions was chosen based on review and consensus among content experts. These items were tested internally with a small group of adults and then a larger group of 80 individuals. From these results, items were changed and reworded so that they could be evaluated on a larger scale in this study.

### Study Participants

Exploratory and confirmatory factor analyses of the VIBE were then conducted with two independent convenience samples. Participants for the exploratory factors analyses (EFA) were recruited from the paid crowdsourcing service Amazon Mechanical Turk (MTurk) while a second sample for the confirmatory factor analyses (CFA) was recruited to complete an online survey through social media, email, word-of-mouth, and the Question Pro website. Respondents completed the rating scales with no personally identifiable information, although optional contact information was collected and stored in a separate file for those interested in a raffle drawing or in participating in future research. The study protocol was approved by commercial vendor Solutions Institutional Review Board (IRB).

The program for completing the rating scales required a respondent to answer an item before moving to the next one, although they could terminate their participation at any time. If two responses were obtained from the same IP address, one was removed. Of note, our data collection occurred in the winter of 2021 while the COVID pandemic was still causing high rates of new cases, hospitalizations, and death.

### The VIBE

The initial 42-item and the final 23-item VIBE consists of a number of statements to which the respondent rates their level of agreement on a 1 to 5 scale as follows: 1) very untrue; 2) somewhat untrue; 3) neither true nor untrue; 4) somewhat true; and 5) very true. The statements used in the VIBE were distributed so that items that were hypothesized to load onto the same dimension were distributed evenly throughout the instrument and not clumped together. Some items were reversed scored so that higher levels of agreement reflected less affinity, rather than more, to a specific dimension, although only one reversed scored item, namely “I’m able to deal with my stress” was retained in the final 23-item version. Respondents who complete the VIBE were asked to rate how they “think, feel, and act most of the time” without mention of a specific timeframe.

### Additional Measures

In addition to the VIBE, the following rating scales were also administered for validity analyses.

#### Personality Traits

The Big Five Inventory (BFI) is a 44-item self-report personality scale based on the Five-Factor Model of personality.^16,17^ Participants rate their agreement to each item along a 5-point Likert scale. The scale assesses five broad dimensions of personality: neuroticism, extraversion, openness, agreeableness, and consciousness. The trait of neuroticism refers to the degree of emotional sensitivity and a predisposition toward experiencing negative emotions like anxiety and sadness. Extraversion encompasses aspects such as activity level, being more outgoing, and enjoyment with being around other people. The trait of openness relates to being creative and receptive to new experiences and ideas. Agreeableness refers to a person being more pleasant, compassionate, and being willing to cooperate in groups to avoid conflict. The trait of conscientiousness, finally, refers to someone’s level of goal orientation, regulatory abilities, and responsibility. The BFI has been shown to have good psychometric properties and strong associations with longer personality instruments that utilize a similar Big Five framework.^16^

#### Depressive Symptoms

To examine depressive symptoms, subjects completed the Patient Health Questionnaire-8 (PHQ-8), an adapted version of the widely used and well-validated screening tool, PHQ-9.^22,23^ The total summary score was used as a quantitative measure of depressive symptomatology. Due to the de-identified manner in which our data were obtained, as well as a lack of clinical backup for positive results, we omitted the item related to suicide and summed the score of the remaining 8 items. Removing this item has not been found to diminish the good validity or reliability of the test.^24^

#### Stress

Subjective stress levels were assessed using the Perceived Stress Scale (PSS), which is one of the most commonly used instruments to measure the perception of stress and has been shown to have good reliability and validity.^25.26^ For the 10-item PSS, respondents indicate on a scale of 0 to 4 how often in the past month they have experienced various types of stress. These items are summed to create an overall stress score.

#### Well-Being

The short version of the Warwick-Edinburgh Mental Well-Being Scale (WEMWBS) was used to assess overall well-being. This instrument has been found to have strong psychometric properties from a number of studies.^27^ It contains 7 positively-worded items in areas such as feeling useful and thinking clearly.^28,29^ Respondents rate how often they experience these items on a 1 to 5 scale and a total sum score is calculated.

### Statistical Analysis

The initial 42 items of the VIBE were first exposed to exploratory factor analysis (EFA). Due to potential problems of overfitting and inflated fit indices, using the same sample for exploratory and confirmatory analyses is not recommended (Fokkema & Greiff, 2017).^30^ Consequently, EFA was conducted using data from respondents recruited through MTurk and confirmatory factor analyses (CFA) were conducted using the data from respondents recruited through the internet and non-MTurk strategies. Maximum likelihood-based EFA with varimax rotation was used explore the factor structure and each item’s loading on resulting factors.^31,32^ The R computing environment package Psych was used to carry out the analyses.^33,34^ The number of factors to be used was based on a number of metrics including eigenvalues > 1 and examination of the scree plot, cumulative variance explained, and interpretability of the model.^35^

Based on the results of the EFA, items were assigned to factors, with unnecessary or poorly loading items removed. The final structure was then subjected to CFA using the sample recruited from non-MTurk strategies and the acceptability of the model was examined using a number of standard goodness-of-fit indices including Cronbach’s alpha coefficient, the average correlation within a scale (also known as alpha 1), the correlation of each item with each scale, the simple signal to noise ratio (i.e., n·r/(1−r) where r is the average item-specific correlation for a factor), and the intercorrelation of all the scales, again using the Psych package in the R environment.

We subsequently attempted to identify clusters of similarly responding subjects on the VIBE using K-means.^36^ While there are many clustering techniques, we used K-means because this method allows us to specify the number of clusters, and optimize it. The optimal number of clusters was found using a grid search with a cost metric based on the increase in variance in the silhouette score. Using the cluster indices as labels, a confusion matrix was created that quantified the discriminability between the clusters of subjects across a range of classifiers (linear discriminating analysis, K-nearest neighbors and linear support vector machine). The VIBE questions were then ranked based on recursive feature elimination using the average discrimination error as a cost metric.^37^

Finally, convergent validity of the final version of the VIBE was tested by calculating correlation coefficients with widely used and validated measures of personality, stress, mood, and well-being, as described above.

## RESULTS

### Demographic Information

As mentioned, EFA and CFA analyses were conducted in a training and test sample. The demographic information for these samples and the overall participant group is shown in Table 1. Almost half (45.6%) of the total sample of 5512 individuals were young adults between the ages of 18 and 24. Regarding gender, a total of 61.4% of the total sample identified as female while 3.6% reported a non-binary or genderqueer identity. Slightly over 10% of the sample came from outside of the United States.

**Table 1.** Demographic information

|  | Training Sample | Test Sample | Total |
| --- | --- | --- | --- |
|  | (N=2049) | (N=3463) | (N=5512) |
|  | N (%) | N (%) | N (%) |
| Age Group |  |  |  |
| 12-17 | 0 (0%) | 885 (25.6%) | 885 (16.1%) |
| 18-24 | 211 (10.3%) | 2297 (66.3%) | 2508 (45.6%) |
| 25-34 | 831(40.6%) | 217 (6.3%) | 1048 (19.0%) |
| 35-49 | 673 (32.8%) | 41 (1.2%) | 714 (13.0%) |
| 50 and above | 334 (16.3%) | 23 (0.7%) | 357 (6.5%) |
| Sex |  |  |  |
| Female | 982 (47.9%) | 2484 (71.7%) | 3466 (62.9%) |
| Male | 1059 (51.7%) | 926 (26.7%) | 1985 (36.0%) |
| No answer | 8 (0.4%) | 53 (1.5%) | 61 (1.1%) |
| Gender |  |  |  |
| Female | 971 (47.3%) | 2412 (69.7%) | 3383 (61.4%) |
| Male | 1049 (51.2%) | 880 (25.4%) | 1929 (35.0%) |
| Genderqueer | 29 (1.4%) | 171 (4.9%) | 200 (3.6%) |
| Trans female | 0 (0%) | 0 (0%) | 0 (0%) |
| Trans male | 0 (0%) | 0 (0%) | 0 (0%) |
| Other | 0 (0%) | 0 (0%) | 0 (0%) |
| No answer | 0 (0%) | 0 (0%) | 0 (0%) |
| Country |  |  |  |
| United States | 2000 (97.6%) | 2889 (83.4%) | 4889 (88.7%) |
| Great Britain | 7 (0.3%) | 158 (4.6%) | 165 (3.0%) |
| Canada | 1 (0.0%) | 66 (1.9%) | 67 (1.2%) |
| India | 7 (0.3%) | 26 (0.8%) | 33 (0.6%) |
| Other | 34 (1.7%) | 324 (9.4%) | 358 (6.5%) |
| Education |  |  |  |
| No HS diploma | 7 (0.3%) | 555 (16.0%) | 562 (10.2%) |
| HS diploma/GED | 438 (21.4%) | 1662 (48.0%) | 2100 (38.1%) |
| Associate's | 187 (9.1%) | 253 (7.3%) | 440 (8.0%) |
| Bachelor's | 960 (46.9%) | 357 (10.3%) | 1317 (23.9%) |
| Masters | 402 (19.6%) | 147 (4.2%) | 549 (10.0%) |
| Doctorate | 48 (2.3%) | 96 (2.8%) | 144 (2.6%) |
| Other | 1 (0.0%) | 202 (5.8%) | 203 (3.7%) |
| No answer | 6 (0.3%) | 191 (5.5%) | 197 (3.6%) |
| Race* |  |  |  |
| White | 1486 (72.5%) | 1849 (53.4%) | 3335 (60.5%) |
| Black AA | 300 (14.6%) | 859 (24.8%) | 1159 (21.0%) |
| Asian American | 216 (10.5%) | 297 (8.6%) | 513 (9.3%) |
| Native American | 34 (1.7%) | 142 (4.1%) | 176 (3.2%) |
| Pacific Islander | 7 (0.3%) | 38 (1.1%) | 45 (0.8%) |
| Hispanic/Latinx | 101 (4.9%) | 670 (19.3%) | 771 (14.0%) |
| Other | 11 (0.5%) | 123 (3.6%) | 134 (2.4%) |
| Don't know | 2 (0.0%) | 33 (1.0%) | 35 (0.6%) |
| No Answer | 9 (0.4%) | 50 (1.4%) | 59 (1.1%) |

HS=High school; GED=General Education Development; AA=African American
\*Ns for race data exceed sample size as respondents were able to select more than one category.

### Exploratory Analyses

Based on a scree plot using the training set data (i.e., the respondents recruited through the MTurk mechanism), the EFA suggested that that VIBE items were best represented by four main factors. Eigenvalues associated with these four factors were 6.72, 5.31, 2.60, and 1.22, respectively, and the total amount of variance across the items explained by the 4 factors was 37%. Upon inspection, the first dimension included items related to the tendency to experience worry and stress and also elements of low confidence. A second dimension was focused around risk taking and enjoyment of high stimulation activities. The third factor included many aspects of valuing relationships and expressing oneself, while the final fourth factor related to goal directed behavior and being deliberate and responsible. These factors were labelled stressed, energetic, social, and disciplined. Items that loaded poorly on these factors (i.e., factor loadings >-0.3 and < 0.3) or loaded too heavily on multiple factors were removed. Table 2 provides the factor loadings of the final 23-item version of the VIBE.

**Table 2.** Factor Loadings from Exploratory Factor Analysis

| Items* | Stressed | Energetic | Social | Disciplined |
| --- | --- | --- | --- | --- |
| 1. I stress out easily. | **0.80** | 0.06 | 0.02 | 0.03 |
| 9. I almost constantly need to check social media on my phone. | **0.38** | **0.33** | .28 | -.08 |
| 18. I have a tough time shaking off negative comments or interactions. | **0.73** | -0.04 | 0.02 | 0 |
| 19. I get less sleep than others. | **0.37** | 0.23 | -0.06 | 0 |
| 26. I worry often and a lot. | **0.81** | 0.01 | -0.04 | 0.07 |
| 33. I lose my cool. | **0.55** | 0.26 | 0.04 | -0.22 |
| 38. I have a lot of doubts about my ability to be successful. | **0.68** | -0.01 | -0.03 | -0.22 |
| 42. I'm able to deal with my stress. (Reversed) | **-0.54** | 0.21 | 0.21 | 0.27 |
| 13. I really enjoy getting "out of my comfort zone." | -0.14 | **0.63** | **0.38** | -0.12 |
| 14. I've purposely taken some big risks in my life. | -0.01 | **0.59** | 0.15 | -0.08 |
| 16. I can get quite competitive. | -0.03 | **0.5** | 0.1 | 0.11 |
| 21. My style and dress draw attention. | 0.08 | **0.52** | **0.36** | -0.16 |
| 27. I seek out opportunities that give me an adrenaline rush. | 0.06 | **0.68** | 0.25 | -0.15 |
| 4. I tend to know the names of lots of service people I interact with regularly (café or cafeteria workers, security staff, bartenders, etc.). | -0.07 | **0.33** | **0.46** | 0 |
| 17. People can usually tell how I feel. | 0.02 | 0.1 | **0.47** | 0.11 |
| 20. I reach out to others when I am upset. | -0.01 | 0.13 | **0.56** | 0.1 |
| 28. I like to show my affection. | -0.03 | 0.1 | **0.63** | 0.17 |
| 34. People are generally trustworthy and kind. | -0.18 | 0.05 | **0.5** | 0.01 |
| 5. I feel uneasy when pushed to break rules and policies. | 0.15 | -0.24 | 0.11 | **0.38** |
| 7. When I run into a problem, I keep at it until it's solved. | -0.26 | 0.16 | 0.14 | **0.57** |
| 15. I carefully consider the options before making big decisions. | -0.07 | -0.08 | 0.06 | **0.53** |
| 32. I set specific goals for myself. | -0.21 | 0.26 | 0.22 | **0.49** |
| 39. I'm rarely late to classes, meetings, or appointments. | -0.12 | -0.06 | -0.01 | **0.4** |

Loadings above 0.30 are shown in **bold**.

To explore the reliability of the four-factor solution, we also pursued other ways of determining the optimal number of factors. Parallel analysis^38^ suggested a 5-factor solution while Velicer’s minimal average partial (MAP) test^39^ suggested 7-factors. However, these additional factor structures beyond the four-factor solution found optimal from the eigenstructure analysis were hard to interpret and only included 1 or 2 items that loaded above 0.3 on those factors. In addition, many items exhibited loadings greater than 0.3 on multiple factors. Thus, overall, due to the coherence and interpretability of the initial four-factor solution, the eigenstructure/scree plot analysis, and the complementary cluster analysis (see below), we settled on this - factor solution.

### Confirmatory Analyses

#### Confirmatory factor analysis

The reliability of the final 23-item version of the VIBE was then explored using a different sample, specifically respondents recruited with methods outside the MTurk mechanism, using CFA techniques. The CFA focused on the individual factors loadings (>0.3 or <-0.3), the internal consistency of the items (i.e., their correlations with the factors and other items contributing to a factor – see Methods) and the results of reliability metrics.^40^ Table 3 suggests that the VIBE four factors have properties in the non-MTurk confirmatory data set consistent with a four-factor solution.

**Table 3.** Confirmatory factor analysis fit indices and factor loadings

| **Reliability Metric** | **Scale** |  |  |  |
| --- | --- | --- | --- | --- |
|  | **Stressed** | **Energetic** | **Social** | **Disciplined** |
| Chronbach's alpha | 0.75 | 0.67 | 0.62 | 0.63 |
| Average correlation | 0.27 | 0.29 | 0.25 | 0.25 |
| Median correlation | 0.27 | 0.28 | 0.24 | 0.24 |
| Guttman's lambda 6 | 0.76 | 0.66 | 0.61 | 0.61 |
| Ratio of intra-/inter-scale correlations | 3.00 | 2.00 | 1.60 | 1.70 |
| **Correlation Matrix** |  |  |  |  |
| Stressed | 0.75 | 0.13 | 0.04 | 0.24 |
| Energetic | 0.09 | 0.67 | 0.72 | 0.47 |
| Social | 0.03 | 0.46 | 0.62 | 0.62 |
| Disciplined | 0.17 | 0.31 | 0.39 | 0.63 |

#### Clustering and Discrimination

The clustering procedure found four clusters of subjects’ responses (scores) to the 23 VIBE questions. The mean of each cluster, corresponding to a column in the heat map, indicates a stereotypical response to the 23 questions. As shown in Figure 1, we found that some subjects (second cluster; second column in the heat map) responded strongly to all questions, while other subjects (third cluster; third column in the heat map) had average responses across all questions. The remaining subjects fell into two categories with generally opposite responses to the same questions. This alternating response pattern highlights how strongly the 23 VIBE questions can elicit divergent responses within a broad audience.

**Figure 1.**
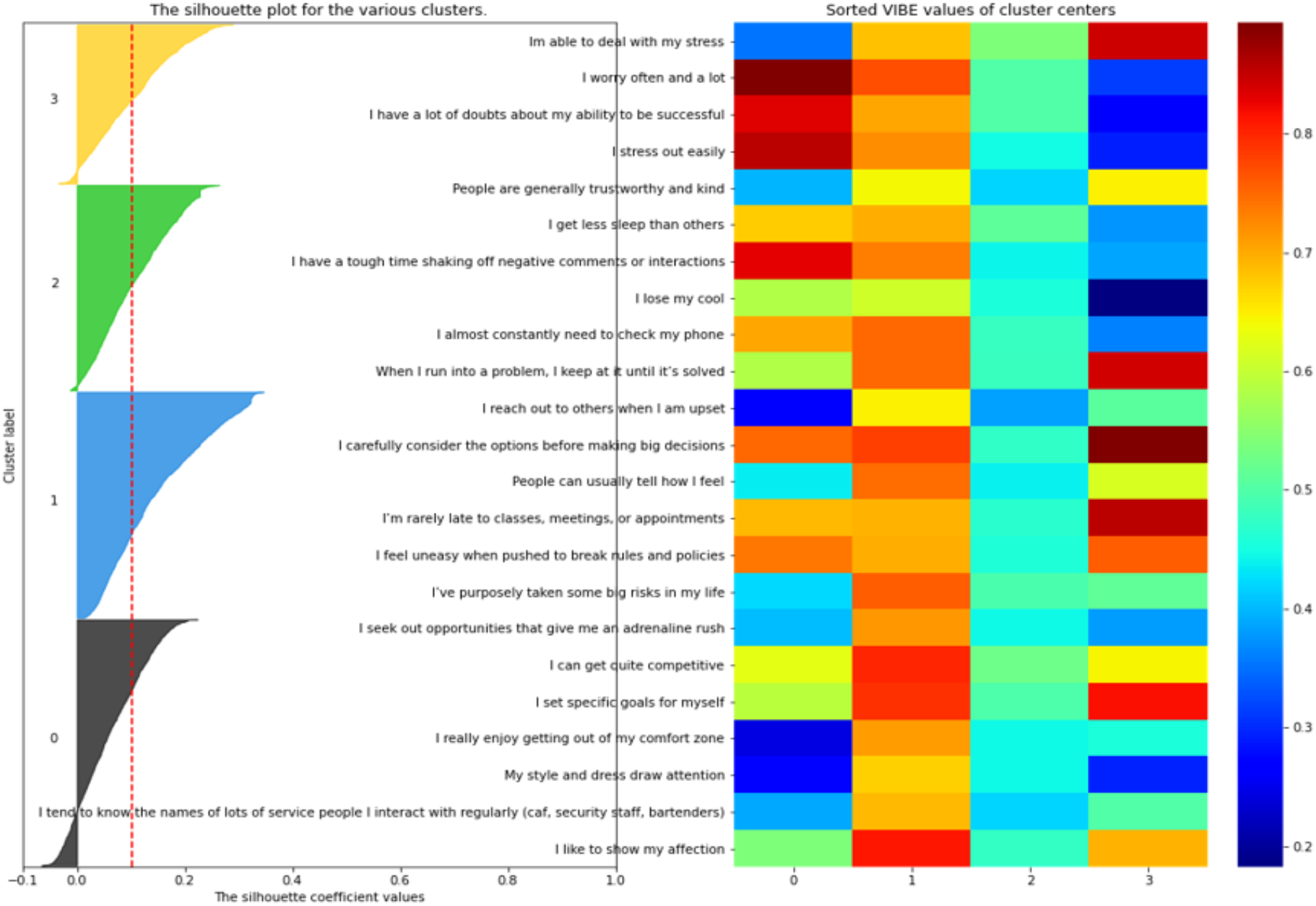
Silhouette analysis for K-means clustering revealing four groups with similar levels of responding to the VIBE. Associated heat map represents the mean of each cluster with red color indicating greater endorsement of that item and blue representing less endorsement.

As shown in Figure 2, the confusion matrix revealed that subjects could be correctly classified into one of these four groups with an average accuracy of 95%. These results corroborate that the 23 VIBE questions intrinsically create four groups of subjects that respond differently to the questions.

**Figure 2.**
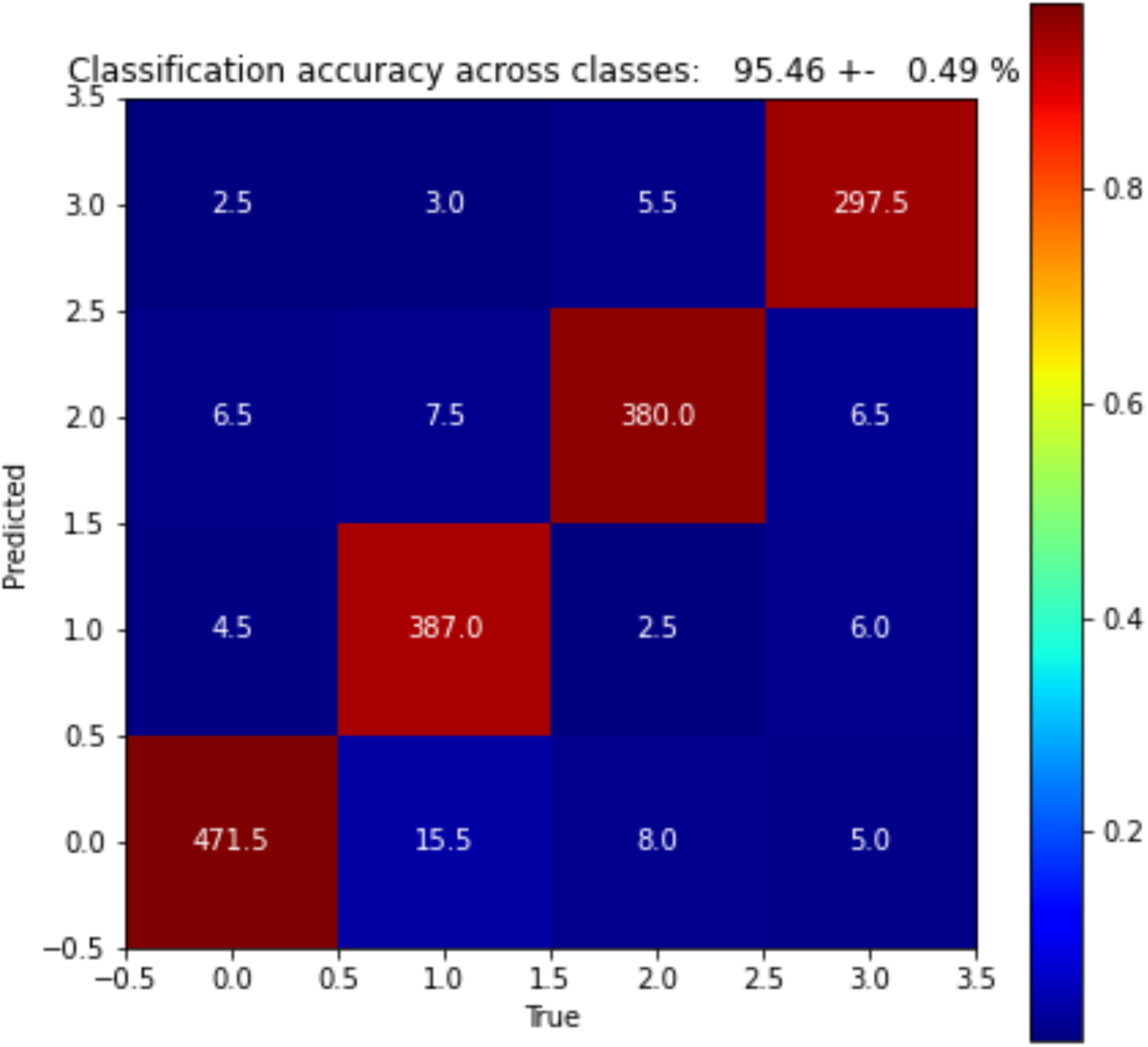
Confusion matrix showing how well the responses to the 23 VIBE questions from the four groups of subjects can be discriminated. The numbers indicate the number of samples, and the color represents the probability across predictions (columns).

**Figure 3.**
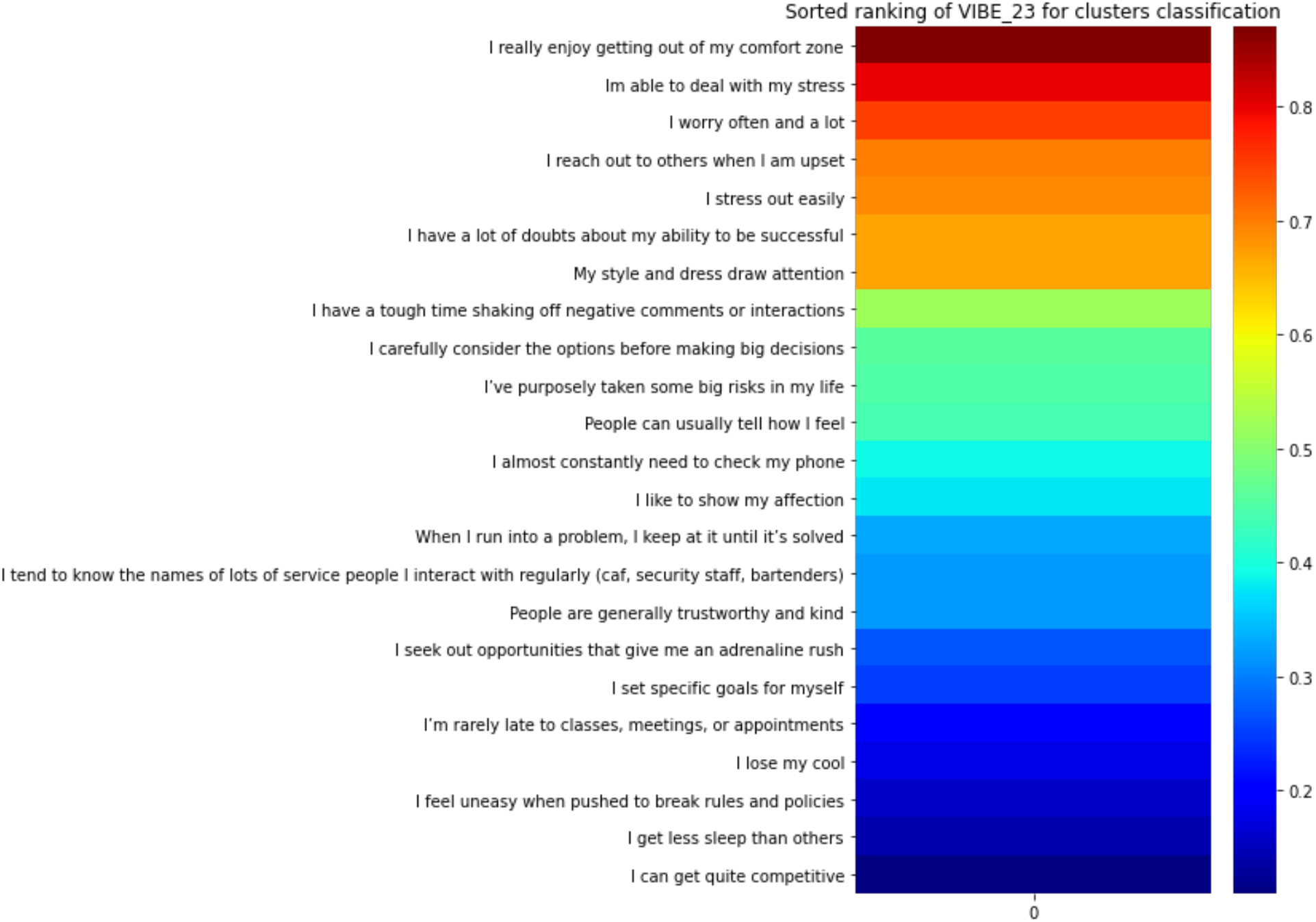
Ranking of VIBE items according to ability to distinguish among the four clusters based on Recursive Feature Elimination and the average classification accuracy across Linear Discriminating Analysis, K-nearest neighbor and linear Support Vector Machine. Red indicates higher ability and blue lower.

Ranking the items based on their ability to discriminate between the four groups showed a wide spectrum of discriminative capability between subjects’ responses based on our questions.

### Validity

Convergent validity for the VIBE was tested by examining standard Pearson correlations between the VIBE and a number of established measures of personality, stress, mood, and well-being. These were calculated using the entire combined sample (N=5512). As shown in Table 4, a number of strong associations were found between these measures and corresponding dimensions of the VIBE. Note that with a total sample size of 5512, a correlation as low as 0.04 would be significant at the 5% type I error level with 80% power. Since we conducted 32 tests, the use of a conservative Bonferroni correction to the type I error (i.e., 0.05/32 = 0.002) would suggest a correlation above 0.05 would achieve statistical significance. As can be seen from Table 4, there were many highly significant associations, even after this multiple comparisons correction. The stressed dimension of the VIBE correlation strongly with the neuroticism dimension of the BFI and the total score of the PSS, in addition to lower ratings of mood and well-being according to the PHQ-8 and WEMWBS, respectively. The energetic dimension, as expected, showed significant correlations with both the extraversion and openness dimensions of the BFI as well as positive associations with mood and well-being. The social dimension of the VIBE also showed a robust association with extraversion with more moderate correlations in anticipated directions with other BFI dimensions and indices of mental health. The disciplined factor was highly correlated with the BFI trait of conscientiousness but also showed moderate and significant associations with other BFI traits and with perceived stress, depressed symptoms, and well-being, all in expected directions.

**Table 4.** Correlations between the VIBE dimensions and established measures of personality, stress, mood, and well-being

| Measure | Stressed | Energetic | Social | Disciplined |
| --- | --- | --- | --- | --- |
| Personality (VIBE) |  |  |  |  |
| Stressed | -- |  |  |  |
| Energetic | **0.13** | -- |  |  |
| Social | **-0.08** | **0.42** | -- |  |
| Disciplined | **-0.10** | **0.13** | **0.39** | -- |
| Personality (BFI) |  |  |  |  |
| Neuroticism | **0.78** | -0.05 | **-0.24** | **-0.22** |
| Extraversion | **-0.23** | **0.43** | **0.42** | **0.08** |
| Openness | -0.04 | **0.27** | **0.24** | **0.29** |
| Agreeableness | **-0.27** | **-0.06** | **0.26** | **0.38** |
| Conscientiousness | **-0.49** | -0.05 | **0.24** | **0.55** |
| Stress (PSS) | **0.69** | 0.02 | **-0.23** | **-0.24** |
| Depressed Mood<br>(PHQ-9<sup>^</sup>) | **0.61** | **0.14** | **-0.14** | **-0.27** |
| Well-Being<br>(WEMWBS) | **-0.42** | **0.22** | **0.43** | **0.38** |

Statistically significant correlations ( $p < .001$ )
BFI=Big Five Inventory; PSS=Perceived Stress Scale; PHQ-9= Patient Health Questionnaire 9; WEMWBS=Warwick-Edinburgh Mental Well-Being Scale ^Suicide item removed from this scale

## DISCUSSION

This study introduces a new 23-item stress-related personality scale, the Virtual Inventory of Behavior and Emotions (VIBE), and tests some of its initial psychometric properties using a sample of over 5000 individuals. While many scales already exist that measure personality traits and related constructs, the brevity of the instrument and incorporation of more modern language and behaviors give the VIBE some properties that could add value, particularly for younger adults. Further, the instrument’s emphasis is on one’s propensity towards experiencing stress and may be particularly useful in work with other stress-related assessment tools. Respondents rate their level of agreement on a 5-point scale to items which encompass statements that include behaviors and feelings such as social media checking, getting out of one’s comfort zone, and ease at breaking rules and policies.

Exploratory factor analyses in a sample of 2049 individuals from a larger initial battery of items revealed a four-factor structure. These dimensions were labelled stressed, energetic, social, and disciplined. The stress factor contained items that directly asked about one’s propensity to experience stress and respond to it by ruminating about negative comments and interactions and having doubts about one’s ability to be successful. Interestingly, an item related to not getting enough sleep also loaded onto this dimension. By contrast, the energetic dimension included items related to enjoying high stimulation activities that can carry risk, being competitive, and presenting oneself in a way that draws attention from others. The social factor included statements regarding being more expressive with one’s feelings and relying on others when upset. It also included items related to having a more positive view about other people and knowing the names of individuals with whom someone has occasional contact. Finally, the disciplined dimension contains elements of being more regulated and goal oriented in addition to being somewhat more cautious and respectful of authority.

These four primary dimensions bear resemblance to other factor structures found when developing temperament and personality instruments. For example, comparing the VIBE factors to the well-known “Big Five” theory of personality scales, there are clear associations between the VIBE dimensions of stressed, energetic, and disciplined with neuroticism, extraversion, and conscientiousness. At the same time, there was far from a one-to-one convergence between individual VIBE dimensions and the Big Five factors, which suggests that the instrument may be offering something new. The social dimension on the VIBE, for example, was shown to have more diffuse associations with all of the Big Five factors. Similarly, the openness dimension from the BFI was significantly related to the energetic, social, and disciplined factors of the VIBE. Previous research has found that openness dimension is less reliably detected in samples of younger individuals.^41^ The four-factor structure of the VIBE also has parallels with both the four dimensions of temperament found within the models of Buss and Plomin,^42^ labelled emotionality, activity, sociability, and impulsiveness, and with the four temperament dimensions, novelty seeking, harm avoidance, reward dependence, and persistence, in the model developed by Cloninger and colleagues.^43^

From these exploratory analyses, a final 23-item version of the VIBE was tested using CFA techniques in a separate sample of 3463 individuals, with nearly half between the ages of 18 and 24. These analyses suggested the four VIBE factors were internally consistent and supported the overall structure of the VIBE. Further, machine learning procedures revealed that the VIBE was highly capable of identifying and distinguishing four groups with very different patterns of responses to the items. These results encourage further investigation with other clustering procedures such as latent class analyses that could be used to identify distinct personality types as they relate to the experience and response to stress.

The final VIBE scales included eight items that loaded on the stressed dimension and five items each devoted to the other three dimensions: energetic, social, and disciplined. At 23 items, the VIBE is considerably shorter than most temperament or personality scales. Nevertheless, strong correlations were found between the VIBE dimensions and established measures of personality, perceived stress, mood, and well-being. The correlation, for example, between the stressed scale of the VIBE and neuroticism dimension of the 44-item BFI was 0.79, and all four dimensions of the VIBE were significantly correlated with well-being. These robust associations lend support for the convergent validity of the VIBE instrument.

As previously mentioned, personality traits have been found to be strongly associated with how one experiences feelings of stress.^8,13^ Consistent with the idea that personalities develop in the context of our interactions with life stressors, and that these traits in turn can lead us to choose environments that perpetuate and reify our stress-personalities, we have found that it can be difficult to disentangle the stress we endure from the persons we become. In many ways, we envision the development of personality in the same way that geoscientists envision the process through which coal, under immense pressure, become diamonds.

The results of our analyses need to be understood in the light of some of our study’s strengths and limitations. While our initial studies with the VIBE were conducted with a large sample with good racial and geographic diversity, it needs to be tested with a more representative and rigorously recruited group, particularly in the context of establishing normative VIBE scale values, as the reliability of personality data from sources like MTurk has been questioned.^44^ These two samples also had differences regarding age, sex, gender, educational level, home country, and race which could have affected the results. Additional tests to verify various forms of reliability, such a test-retest, inter-rater, and construct validity also need to be performed to confirm the psychometric integrity and utility of the VIBE instrument. It is also possible that the COVID pandemic led to changes in the baseline levels of the VIBE traits and their relations to other variables. Nonetheless, the VIBE performed well on a number of both traditional and more modern psychometric analyses in a sample of more than 5000 individuals.

In conclusion, the VIBE is a promising new scale to assess personality, particularly with regard to the experience and response to stress. While our initial studies are quite encouraging, further analyses are planned to solidify the scale’s psychometric foundation and to explore its relations to stress and related constructs, such as burnout, in more detail. For example, individual profile-based analyses will be conducted to identify empirically derived personality types who exhibit similar profiles across the four domains. In addition, personality data should be analyzed in conjunction with knowledge of an individual’s typical stress responses and coping mechanisms to gain a fuller picture of how various stress experience profiles may correlate with various stress coping strategies, as well as how effective these strategies are. We are planning studies to examine the relations between stress-based personality types and various physiological markers of stress through wearable technologies (e.g., electrodermal activity monitors). Our ultimate goal is to implement and make available the VIBE and related scales to individuals so that they can obtain personal insights about their own experience and response to stress. This may empower them to develop strategies and make choices that can build their resilience and allow stress, at least in moderate levels, to function as a growth-promoting rather than growth-inhibiting component of their lives.

## Data Availability

Data from this study are not available due to the raw data being important intellectual property for Happy Health, Inc.

